# Assessing causal relationships between oral microbiota and Periodontitis: evidence from Mendelian randomization analysis

**DOI:** 10.64898/2026.02.01.26345317

**Authors:** Zi-feng Wei, Jia-pei Wuzhang, Yi-ting Huang

## Abstract

**Objective:** This study investigates potential causal relationships between the oral microbiome and periodontitis in East Asian populations, aiming to identify microbial taxa involved in disease pathogenesis.

**Methods:** We performed a two-sample Mendelian randomization (MR) analysis using genome-wide association study (GWAS) summary statistics for tongue dorsum and salivary microbiomes alongside periodontitis data in East Asian populations. Causal estimates were primarily obtained using the inverse-variance weighted (IVW) method, with MR-Egger, weighted median, weighted mode, and simple mode approaches serving as complementary analyses. Sensitivity analyses, including Cochran’s Q test, MR-Egger intercept test, MR-PRESSO, and Steiger filtering, were conducted to assess heterogeneity, horizontal pleiotropy, and reverse causality.

**Results:** At species-level resolution, 60 microbial taxa exhibited evidence of causal association with periodontitis. Of these, 29 were negatively associated and 31 showed a positive association. These taxa were predominantly enriched in the genera *Campylobacter_A, Pauljensenia, Solobacterium*, and *Streptococcus*.

**Conclusion:** This study provides preliminary genetic evidence for causal links between specific species-level oral microbial taxa and periodontitis. Given the limited sample size, these findings should be interpreted as hypothesis-generating and require confirmation in larger independent cohorts.

## 1. Introduction

Periodontitis is a chronic inflammatory disease initiated by dental plaque biofilms. It represents the leading cause of tooth loss in adults and contributes to the etiology of multiple systemic conditions [1]. Dysbiosis of the oral microbiome is considered a central driver of periodontitis pathogenesis. While observational studies have consistently correlated specific microbial abundances with disease status, such associations are susceptible to environmental confounding and reverse causation, impeding definitive causal interpretation.

MR offers a robust framework to strengthen causal inference by leveraging genetic variants as instrumental variables (IVs). Recent GWASs of the oral microbiome in East Asian populations have identified robust genetic instruments for microbial taxa. However, GWAS data for periodontitis in East Asian ancestry remain limited in sample size. The primary publicly available dataset is derived from East Asian immigrants in the United States, which introduces potential concerns regarding environmental confounding and generalizability.

Notably, published MR studies on oral microbiota and periodontitis have inappropriately paired East Asian exposure data with Finnish-ancestry outcome data. According to the STROBE-MR guidelines, such cross-ancestry designs violate the “same underlying population” assumption of two-sample MR, introducing attenuation bias that underestimates the true causal effects [2, 3]. This concern has been corroborated by multiple genetic studies [4-7].

In this context, we conducted an exploratory two-sample MR analysis to investigate whether genetically predicted abundances of oral microbial taxa causally influence periodontitis risk in East Asians. Given the modest case count in the outcome GWAS, our findings warrant cautious interpretation.

## 2. Methods

### 2.1. Study Design and Data Sources

This study adhered to the STROBE-MR guidelines [2, 3]. We employed a two-sample MR framework utilizing summary statistics from GWASs of East Asian populations, covering 3,117 oral microbiome traits and periodontitis (Figure 1). The analysis was grounded in the three core assumptions of MR: (1) the genetic variants are robustly associated with the exposure (oral microbiota); (2) the variants are not associated with confounders of the exposure-outcome relationship; and (3) the variants influence periodontitis solely through the oral microbiome.

**Figure 1.**
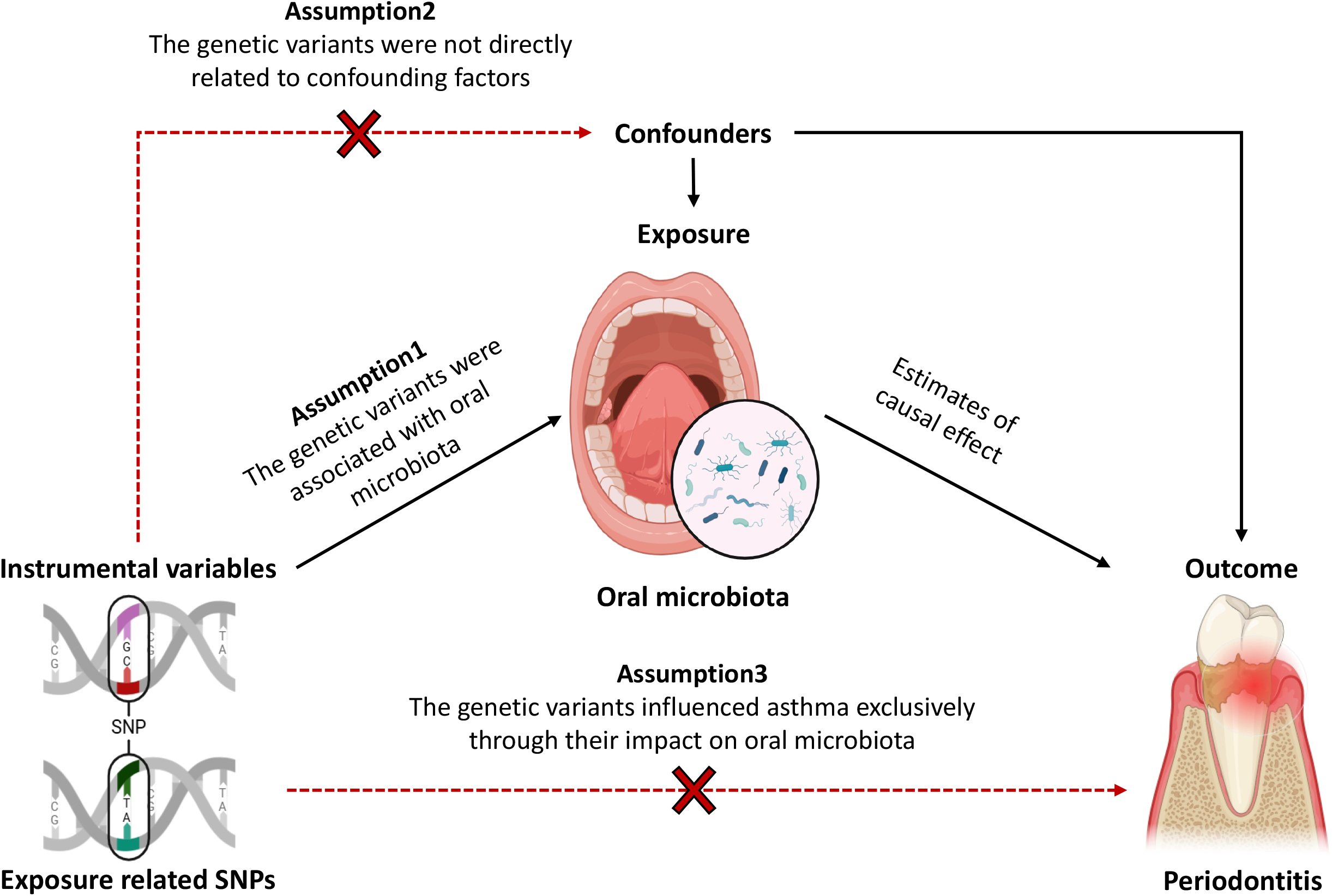
Study design of the MR between oral microbiota and asthma. The solid lines represent the associations between the IVs and oral microbiota, as well as between oral microbiota and the outcome. Dashed lines with a cross indicate the two basic assumptions of MR: the IVs are not directly associated with confounding factors, and the IVs influence asthma exclusively through their effect on oral microbiota.

Oral microbiome data were obtained from a large-scale GWAS available in CNGBdb (CNP0001664), which included 2,017 tongue dorsum samples and 1,915 salivary samples from healthy Chinese individuals [8]. Periodontitis data were sourced from the GWAS Catalog (GCST90478258), comprising 703 cases and 5,762 controls of East Asian ancestry residing in the United States [9, 10].

### 2.2. Instrumental Variable Selection

IVs were selected based on the following criteria: (1) Single nucleotide polymorphisms (SNPs) associated with each oral microbial trait were identified at a genome-wide significance threshold of *p* < 1×10^-5^; (2) Only SNPs with a minor allele frequency (MAF) > 0.01 were retained; (3) To ensure independence, SNPs were clumped to remove linkage disequilibrium (LD) using a threshold of *r*^*2*^ < 0.001 within a 10,000 kb window [11]; (4) The strength of the instruments was validated using the *F*-statistic, calculated as *F = (beta/se)*^*2*^, where SNPs with an *F* < 10 were excluded to mitigate weak instrument bias.

### 2.3. MR Analysis

The IVW method was employed as the primary analysis to estimate the causal effect, with statistical significance set at *p < 0.05*. To ensure the robustness of the estimates, we applied supplementary methods including MR-Egger regression, weighted median, weighted mode, and simple mode. A causal relationship was considered suggestive if the direction of the effect estimate was consistent across the IVW and supplementary methods.

### 2.4. Sensitivity Analysis

Heterogeneity among IVs was assessed using Cochran’s Q test; *p* > 0.05 indicated no significant heterogeneity affecting the IVW results. Horizontal pleiotropy was evaluated using the MR-Egger intercept and the MR-PRESSO global test, where *p* > 0.05 indicated an absence of significant pleiotropy. Finally, Steiger filtering was applied to confirm the directionality of the causal effect and rule out reverse causality.

## 3. Results

The MR results regarding the causal effects of oral microbial taxa on periodontitis are presented in Figure 2 and Figure 3. At the species level, we identified 60 taxa with causal associations with periodontitis. Among these, 29 taxa exhibited a negative association (Odds Ratio [OR] < 1), while 31 taxa showed a positive association (OR > 1). These microbial taxa were predominantly enriched in the genera *Campylobacter_A, Pauljensenia, Solobacterium*, and *Streptococcus* (Figure 4). The Sankey diagram illustrates the associations between microbial taxonomic units (Figure 5).

**Figure 2.**
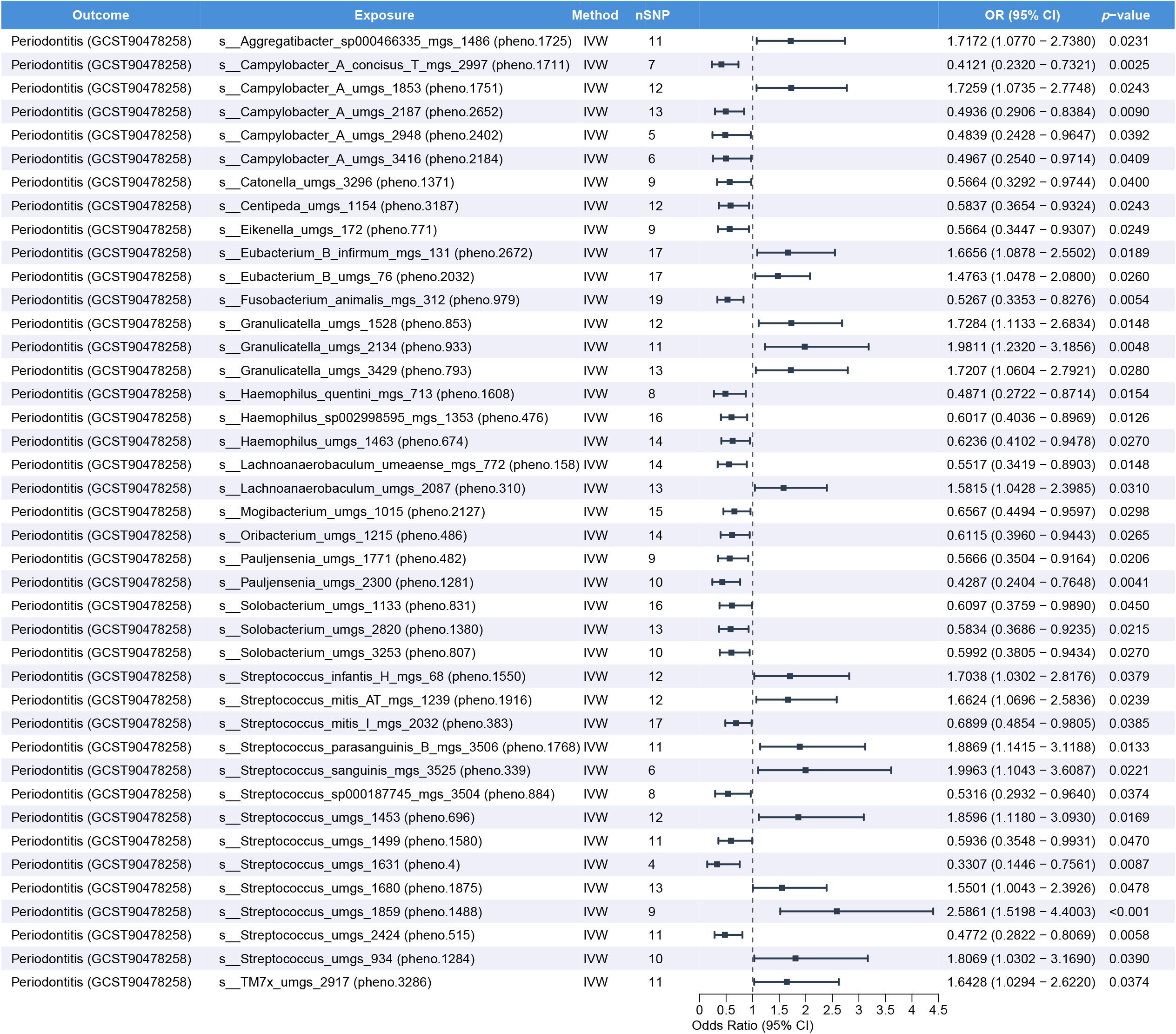
Forest plot showing the causal associations between tongue dorsum microbiota and periodontitis.

**Figure 3.**
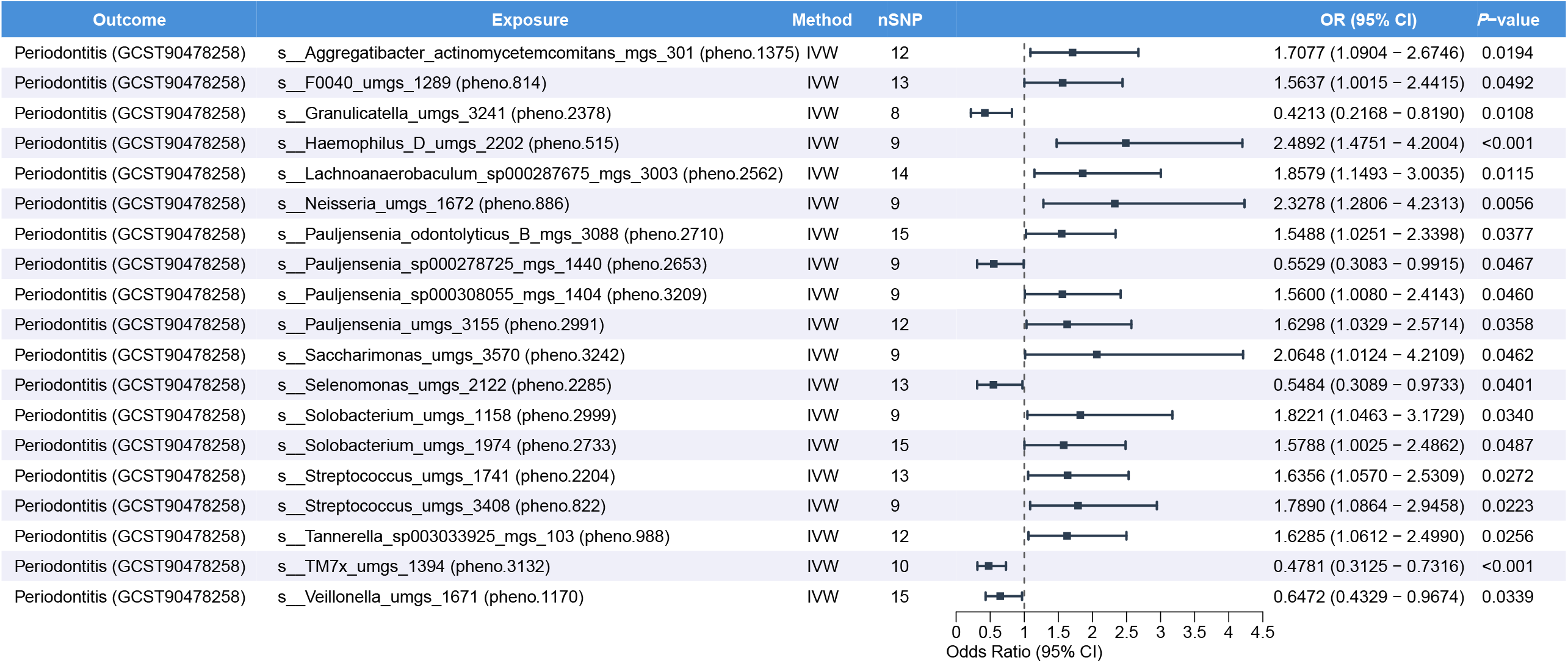
Forest plot showing the causal associations between salivary microbiota and periodontitis.

**Figure 4.**
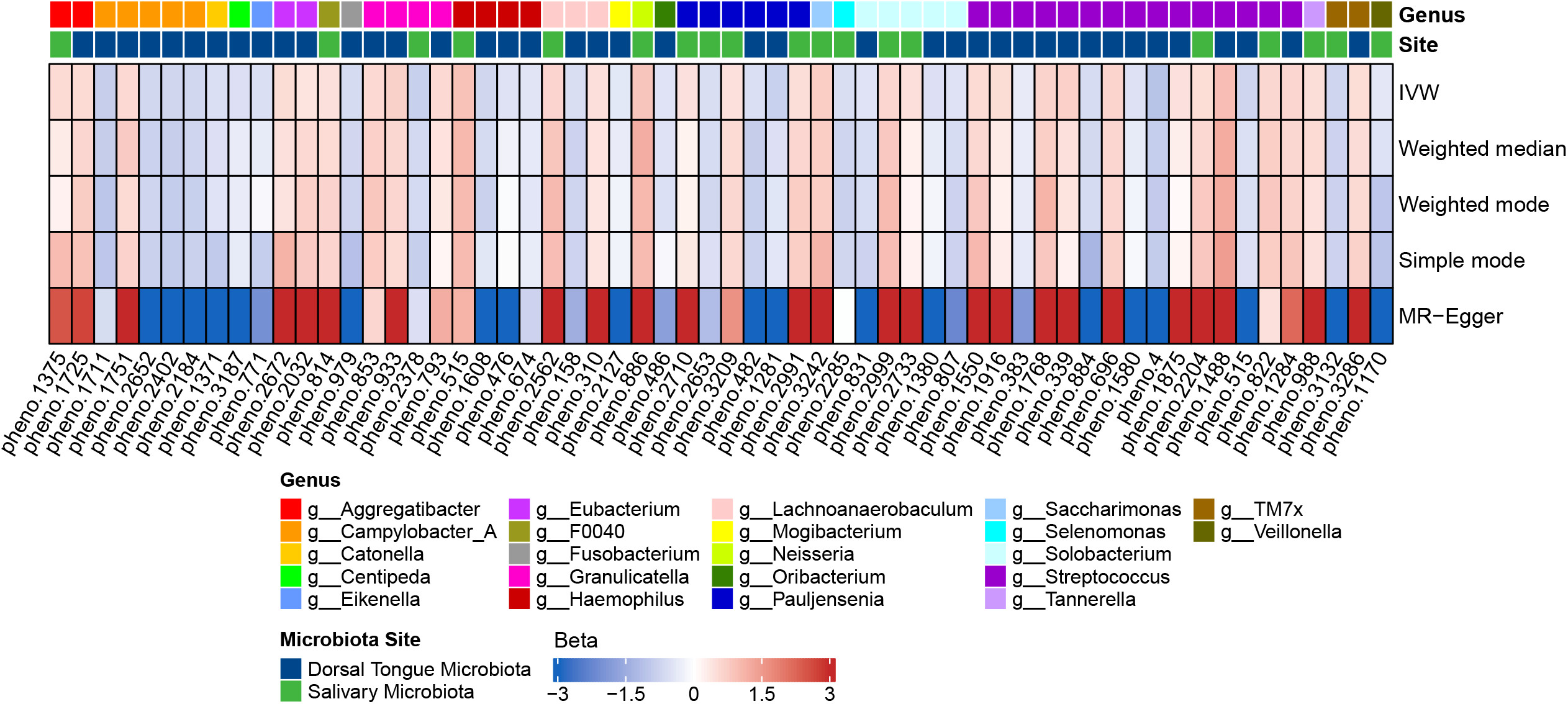
Results of the analysis of oral microbiota causally associated with periodontitis.

**Figure 5.**
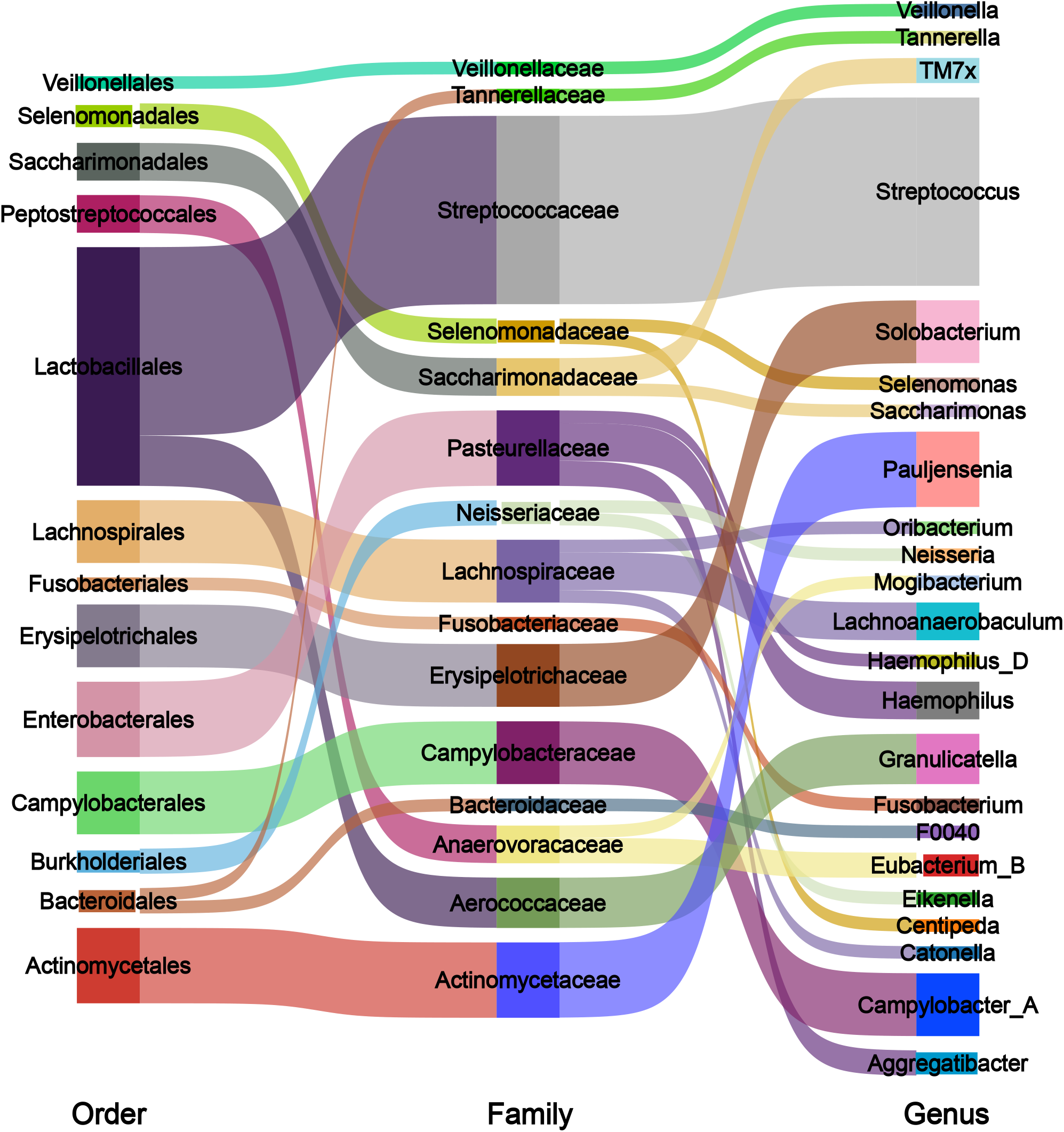
Sankey diagram showing the associations of oral microbiota causally linked to periodontitis at the order, family, and genus levels.

Comprehensive sensitivity analyses confirmed the robustness of the primary estimates. The Cochran’s Q test indicated no significant heterogeneity among the instrumental variables (*p* > 0.05). The MR-Egger intercept test and the MR-PRESSO global test both yielded *p*-values greater than 0.05, suggesting the absence of substantial horizontal pleiotropy. Furthermore, Steiger filtering confirmed that the observed associations were not driven by reverse causation, as the variance explained by the SNPs in the exposure was significantly higher than that in the outcome (*p* < 0.05). Detailed statistics for all MR models and sensitivity tests are provided in Supplementary Tables 1-12.

## 4. Discussion

This study leveraged recent large-scale GWAS data to explore the causal links between the oral microbiome and periodontitis in East Asian populations via a two-sample MR approach.

Several limitations must be acknowledged. First, periodontitis is a heterogeneous disease comprising various subtypes (e.g., chronic versus aggressive), each possessing distinct pathophysiological profiles. Due to the lack of granular phenotypic data in the summary statistics, we were unable to stratify our analysis by disease subtype or severity. Second, we recognize that the oral microbiome is not the sole determinant of periodontitis; postnatal environmental factors and dietary habits play significant roles in disease onset and progression. Third, the statistical power was constrained by the modest sample size of the outcome GWAS, which may limit the precision of effect estimates and increase the risk of false positives. Finally, as the periodontitis GWAS data were derived from East Asian veterans residing in the United States, the generalizability of these findings to diverse East Asian population strata remains to be validated using larger, more representative cohorts from various geographical regions.

## 5. Conclusion

This exploratory two-sample MR analysis provides preliminary evidence for causal effects of specific oral microbial taxa on periodontitis risk in East Asian populations.

## Data Availability

All data analyzed in this study are publicly available. Oral microbiome GWAS summary statistics were obtained from the CNGBdb (Accession: CNP0001664). Periodontitis GWAS summary statistics were sourced from the GWAS Catalog (Accession: GCST90478258). Links to these repositories and the corresponding summary statistics are included within the manuscript and its supplementary materials.

https://ftp2.cngb.org/pub/CNSA/data5/CNP0001664/

## Author contributions

Zi-Feng Wei: Conceptualization, Data curation, Formal analysis, Methodology, Project administration, Software, Supervision, Validation, Visualization, Writing - original draft, Writing - review and editing. Jia-Pei Wuzhang: Writing - review and editing. Yi-Ting Huang: Writing - review and editing.

## Data availability

The GWAS summary data for salivary and dorsal tongue microbiota are sourced from CNP0001664 (CNGB database: https://ftp2.cngb.org/). The GWAS summary data for periodontitis analyzed in the current study are sourced from GCST90478258 (GWAS Catalog: https://www.ebi.ac.uk/gwas/).

## Declarations

### Ethics statement

The present study does not require ethical approval.

